# Organizational Factors Improving Maternal and Newborn Healthcare Quality: Evidence from Ethiopian Health Facilities

**DOI:** 10.1101/2025.10.22.25338601

**Authors:** Abera Biadgo Kefale, Gebremeskel Tamene, Abebe Abraha, Mehiret Abate, Nebiyou Wendwessen Hailemariam, Abiyou Kiflie

**Author notes:** **Corresponding Author:** ABK or.

## Abstract

**Background:** Poor quality maternal and neonatal healthcare continues to contribute to preventable deaths and long-term disabilities in low-resource settings. Organizational factors within health facilities influence the implementation and uptake of evidence-based interventions, affecting health system performance and policy outcomes.

**Objective:** To identify organizational factors that improve the quality of maternal and neonatal healthcare services and examine their implications for evidence-informed decision-making in Ethiopian health facilities.

**Methods:** A facility-based pre–post evaluation was conducted across 131 health facilities in three regions of Ethiopia, implementing a phased package of quality improvement interventions. Mixed-effects Poisson regression was used to assess organizational predictors of improvement in clinical bundles and maternal and neonatal health (MNH) quality indicators.

**Results:** Improvements were observed in at least one clinical bundle in 103 (78.6%) facilities, and in MNH quality indicators in 117 (89.3%) facilities. Significant organizational predictors for clinical bundle improvements included teamwork (IRR = 1.87; 95% CI 1.06–3.29), regular client satisfaction surveys (IRR = 1.27; 95% CI 1.12–1.44), and the presence of a quality structure with clear terms of reference (IRR = 1.34; 95% CI 1.23–1.47). Facilities implementing all three interventions demonstrated the strongest combined effect (IRR = 3.55; 95% CI 2.53–5.00). For MNH quality indicators, predictors included quality improvement knowledge (IRR = 1.34; 95% CI 1.08–1.67) and hospital-level facilities (IRR = 1.35; 95% CI 1.08–1.67), while resistance to change (IRR = 0.73; 95% CI 0.58–0.92) and staff turnover (IRR = 0.62; 95% CI 0.50–0.76) were negatively associated.

**Conclusion:** Teamwork, structured quality systems, and client feedback appear to support improvements in maternal and newborn care. Quality improvement knowledge and hospital-level facilities were associated with MNH quality indicators, while resistance to change and staff turnover showed negative associations. Considering these factors in health system planning may help inform policy, strengthen governance and contribute to better outcomes in low-resource settings.

## Introduction

Maternal and neonatal mortality and morbidity remain major public health challenges worldwide, particularly in low- and middle-income countries (LMICs), including Ethiopia (1, 2). Poor-quality maternal and neonatal healthcare contributes significantly to preventable adverse outcomes and deaths (3). Key challenges include limited access to skilled care, gaps in service delivery and systemic inequalities within health system (4, 5). In Ethiopia, the mean quality scores for obstetric and newborn care have been reported at about 73%, while only 34% of health facilities met the recommended standards (6). Although global initiatives such as the Sustainable Development Goals (SDGs) aim to reduce maternal and neonatal mortality, persistent gaps in service quality continue hamper progress toward achieving the targets (7).

Quality of maternal and newborn healthcare requires safe, effective, and patient-centered, services that meet established standards and improve health outcomes (8-10). Evidence suggests that about 58% of deaths are associated with poor quality of care (8). To evaluate and improve service delivery, health systems increasingly rely on quality indicators, which provide measurable benchmarks for clinical practice and outcomes (8, 9). Monitoring improvements in these indicators allows health facilities and policymakers to identify gaps, track progress, and guide targeted interventions for maternal and newborn care (8, 9).

Improving healthcare quality is a continuous process that involves systemic efforts to strengthen service delivery and health outcomes (11-14). Evidence from quality improvement (QI) initiatives in Ethiopia has shown measurable improvements in maternal and newborn service indicators (6, 15, 16). For instance, the proportion of mothers attending four antenatal care visits increased from 64.1% to 75.3%, syphilis testing during antenatal care increased from 54.7% to 68.5%, and early postnatal care within 48 hours increased from 49.4% to 58.2% following implementation of QI intervention (6, 15, 16). These findings demonstrate that QI initiatives supported by organizational commitment can to measurable improvements in MNH quality indicators and clinical bundle adherence (12, 17-19).

In Ethiopia, several national initiatives have been introduced over the past two decades to improve healthcare quality (20). One of the major frameworks is the National Quality and Safety Strategy (NQSS) (20). This strategy builds on earlier initiatives to strengthen service delivery and improve health outcomes (20-23). The development of the NQSS was a collaborative effort led by the Ministry of Health (MoH), with technical support from the Institute for Healthcare Improvement (IHI) and the involvement of partners and key stakeholders (20). These initiatives emphasize the use of standardized quality indicators and clinical bundles to monitor performance and guide quality improvement efforts (3, 20).

Despite these efforts, variations in the quality of maternal and newborn health services remain a significant challenge (13, 17). These variations are often influenced by organizational factors within health facilities including leadership support, staff capacity, and institutional commitment to quality improvement (13). However, limited evidence exists on how organizational factors influence the implementation effectiveness of QI initiatives and improvements in maternal and newborn health quality indicators. Therefore, this study aims to identify and examine the organizational factors influencing the improvement in MNH quality indicators during the implementation of quality improvement initiatives.

## Methods

### Study area

Ethiopia is the second most populous country in Africa. The country has multiple cultures and languages, with more than 86 across twelve regions and two city administrations. The study was conducted in three regions - Amhara, Oromia, and Southern Nations, Nationalities, and Peoples (SNNP). A total of 131 health facilities were included in six selected zones across the three regions in Ethiopia.

### Project information

The quality initiative Phase II scale-up project was implemented by the Institute for Healthcare Improvement and the Ethiopian Ministry of Health. It was implemented from October 2019 to September 2022 in five regions of Ethiopia (Afar, Amhara, Oromia, SNNP, and Tigray). The project engaged 27 zones, 384 woredas, and 1,670 health facilities (1,505 health centers and 165 hospitals) across three cohorts. The initiative aimed to strengthen health system quality and equity through quality improvement (QI) approaches. It was implemented at the zonal level, with woredas (districts) serving as the scalable unit. Overall, 5,157 health workforces were trained, 2,582 professionals participated in the learning sessions, 3,321 received coaching support, and 2,875 QI projects were implemented across participating facilities.

### Study design

A facility-based pre–post interventional evaluation design was deployed to assess the quality initiative phase II scale-up project.

### Interventions and core components

The intervention package included the following core components:

a. Capacity building: QI training for coaches (2 to 3 per facility, 3 per zone) and basic QI training for frontline care providers (2 per facility)
b. Baseline assessment and continuous monitoring
c. Identifying gaps and prioritizing focus areas
d. Quarterly learning sessions at the woreda and zonal level
e. Regular coaching visits to facilities
f. Capacitating zonal and woreda coaches to provide ongoing coaching support to health facilities
g. On-site orientations to the health facility staff

These activities were delivered in a phased approach, with six months of active support per zone followed by six months of follow-up. During this period, health facilities were expected to implement quality of care activities, including: establishing quality structures, preparing Terms of Reference (ToR), developing and updating quality plans and QI dashboards, designing and implementing QI projects.

### Measurements

In this evaluation, MNH and clinical bundle outcomes were used to measure the quality of MNH care in health facilities. Indicators were selected through an expert panel discussion prior to project implementation, targeting eleven MNH quality and seven clinical bundle indicators.

A bundle consists of three to five clinical care elements that should be provided to all patients every time. For this evaluation, the seven bundle outcome indicators, each included their key bundle elements, and all-or-none principles were applied to monitor progress.

Improved bundle indicators were determined using run chart improvement rules, specifically shift (six consecutive data points above or below the median) and trend (six consecutive points in increasing or decreasing order). When a bundle indicator showed a signal of improvement, it was counted as “improved” for that facility. Similarly, improved MNH quality indicators were counted when run chart rules signalled improvements. It has eleven individual MNH indicators. Facilities showing no improvement from any of the indicators received a count of zero. Analyses were conducted separately for each indicator in a dedicated database, resulting in a count of improved indicators per facility.

These counts of improved indicators were analysed using Poisson regression to identify factors associated with improvements. In this context, the term “incidence” reflects the rate at which improvements occurred across facilities, capturing trends in quality improvement over time rather than absolute static values.

This study is guided by a conceptual framework drawing on the World Health Organization health system innovation and scale-up framework and implementation science concepts. The framework conceptualizes organizational factors as key determinants of QI implementation processes and subsequent improvements in MNH indicators (details in Supplementary File 1).

### Study population

The study population included all medical records of antenatal care, delivery, and complications, and all health care providers and managers working in the selected health facilities.

### Sample Size

Data from the Phase-I work suggest that the intervention would lead to noticeable improvements in most bundle measures. We supposed it was essential to detect at least a 20% absolute improvement in these performance measures. This means, for example, detecting an increase from 75% to 95%, or from 60% to 80% in the proportion of charts meeting specific quality measures.

For the pre-post analysis, we assumed that sampling five charts per measure per facility per month would be reviewed from selected hospitals and health centers in Oromia. This resulted in a sample size of 1470 for each time period. With this sample size, we had 90% power to detect (using a significance level of 0.05) relative improvement in the bundle measures from an average of 75% to 95% within the three zones in Oromia. Similarly, we had 80% power to detect a relative improvement from 60% to 80%. Similar power is available to detect the same range of differences within the other selected zones in the Amhara and SNNP regions. We considered the α-value at 0.05 and 95% level of significance in a two-tailed test.

The unit of analysis was the health facility. Using the two-population proportion sample size calculation formula at a ratio of 1 (baseline phase): 3 (active, follow-up, and sustainability phases), a power of 80%, and a 95% confidence level, we found a sample size of 307 for each of the ANC and delivery-related indicators. Considering stratification by zone and Woreda, we used a design effect of 1.5, which resulted in a sample size of 461 for each of the ANC and delivery indicators. On average, there were four health centers per woreda. Dividing 461 by 4 gives us 115 records needed to be reviewed per indicator per facility. Dividing this number by the total number of months for the study (24) to get 4.8 (approximately 5) per month per facility per indicator.

### Sampling procedure

Stratified random sampling combined with a quota approach was used. The study focused on three regions from the second cohort, which had a similar proportion of zones and received a comparable intervention package and timeframe. Within each region, zones were proportionally allocated, and 33% of zones were randomly selected. A total of 56 woredas were included. These were randomly selected from the chosen zones, with 50% of woredas in each zone sampled. Health facilities were sampled using a quota approach: 50% of hospitals and 20% of health centers were randomly selected.

As shown in Fig. 1, zonal, woreda, and facility lists, along with antenatal care and delivery registers, were used as the sampling frame. Systematic random sampling was then applied to select medical charts.

**Fig 1:**
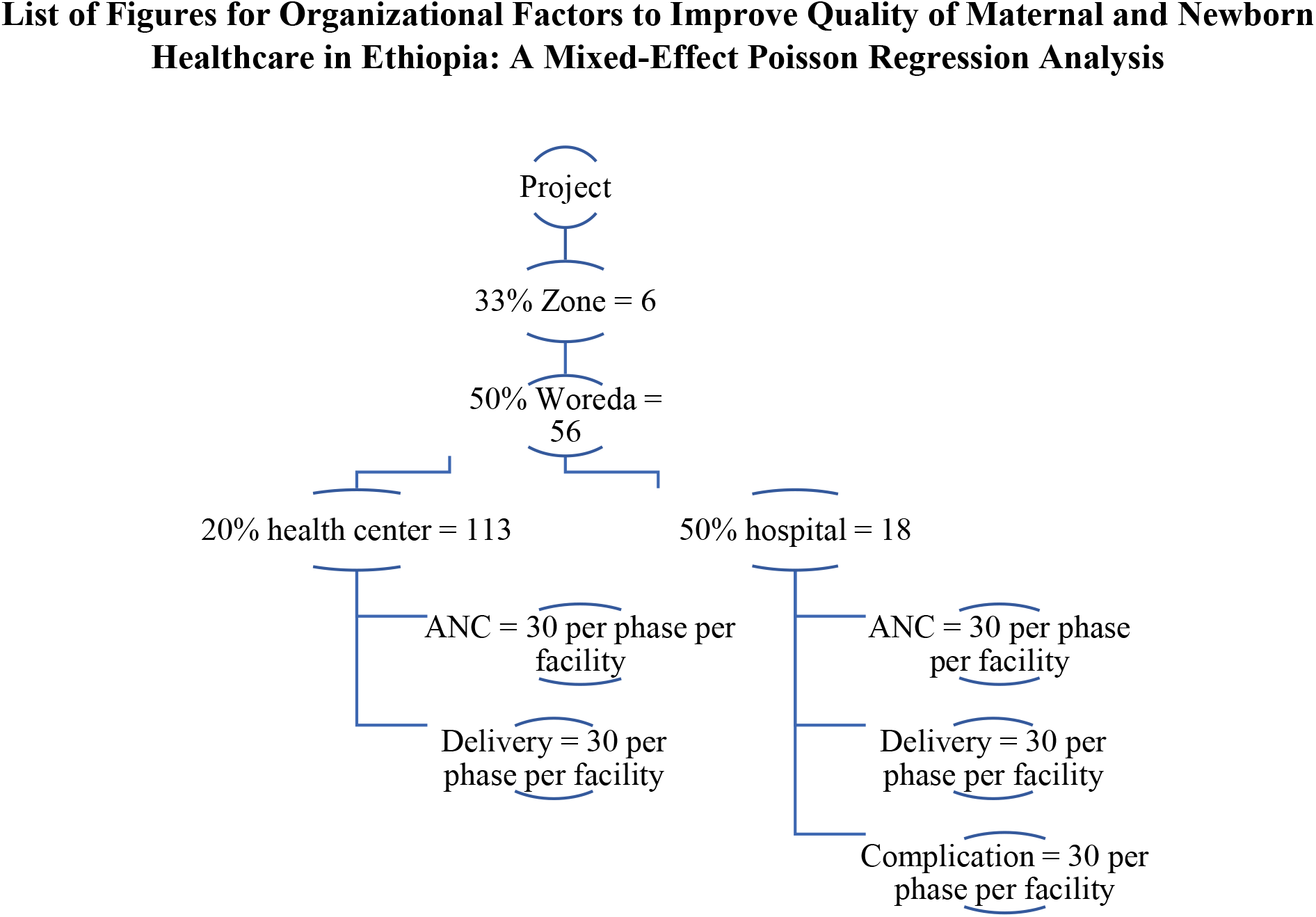
Sampling procedure for Ethiopia Health care quality initiative evaluation

### Data collection tools and techniques

Health facilities are working to improve the health system to achieve the intended performance on maternal and newborn health outcomes, as measured by standard indicators. Some indicators are set and regularly collected, entered, and transmitted through electronic-based MNH; however, most healthcare data, in particular care process data, are not regularly collected and analysed. Therefore, to measure clinical processes, the national quality initiative scale-up project used clinical bundle indicators and selected key MNH quality indicators to assess the implementation effectiveness.

Data were collected from health facility heads and frontline providers through interviews and direct observations of facility structures, QI activities, and supporting documents. These methods captured antecedents, leadership engagement, staff engagement, and other QI-related variables. Additional sources included patient charts, facility reports, QI documentation, and project office archives. Data access occurred from 17/09/2021 to 13/10/2021 and 10/02/2022 to 31/02/2022, with no individual participants identifiable.

To ensure data validity and reliability, data collectors and supervisors were trained in the objective, purpose, and methods of data collection. The data collectors and supervisors were health professionals who had experience in quality improvement activities in the health sector. The data collection tools were tested in 10 of the sampled health facilities. Moreover, the evaluation data were carefully handled and daily checked for cleanliness, completeness, and consistency by supervisors during data collection with the principal investigator’s guidance.

### Data analysis

The data were checked for completeness, consistency, and clarity accordingly. Data were entered into an Excel spreadsheet, and the relevant variables for this paper were extracted, coded, and exported to Stata version 17 for analysis. Descriptive statistics were computed to determine the frequency distribution of the responses, which is presented either in tabular or graphical form. Run chart rules were applied to determine whether the selected MNH quality indicators showed a signal of improvement. Then, the number of improved MNH quality indicators was summarized by each health facility. A mixed-effects Poisson regression analysis was used for MNH clinical bundle indicators, and a Poisson regression analysis was used for MNH quality indicators to identify potential predictors. P-value <0.05 was considered statistically significant.

#### Mixed-effect Poisson regression analysis method

Model fitness checking and overdispersion testing had been done; no overdispersion has been observed. Clinical bundle indicators (observed variance of 1.87 and a mean of 2.02, which makes a variance-to-mean ratio of 1.08. No excess zeros were detected. Moreover, model goodness-of-fit was evaluated. Akaike information criterion (AIC) and Bayesian Information Criterion (BIC) were used to assess model fitness and suitability. Further cluster effects were measured using the Intraclass Correlation Coefficient (ICC) measurement (Table 1).

The null model (model-1) was fitted without any explanatory variables. A random-intercept Poisson regression analysis (model-2) was then fitted to estimate associations between individual independent variables and the number of improved MNH clinical bundle indicators using the ‘mepoisson’ command in STATA. Then, the model (model-3) was adjusted to assess effect modification in the association of predictor variables and the number of improved MNH clinical bundle indicators. Theoretical and iterative statistical methods, in addition to forward selection and backward elimination processes, were applied to identify explanatory variables.

#### Poisson regression analysis method

Model fitness checking and overdispersion testing have been done; no overdispersion has been observed. MNH quality indicators (observed variance of 2.31 and a mean of 2.35, which makes a variance-to-mean ratio of 1.02. No excess zeros were detected. Moreover, model goodness-of-fit was evaluated. Akaike information criteria (AIC) and Bayesian Information Criteria (BIC) were applied to assess model fitness and suitability (Table 1). Additionally, multicollinearity was checked for model fitness, and none was detected.

The null model was fitted without any explanatory variables. A Poisson regression analysis was then fitted to estimate associations between individual independent variables and the number of improved MNH quality indicators using the ‘poisson’ command in STATA. Then, the multivariable model was adjusted to assess the association of predictor variables and the number of improved MNH quality indicators. Theoretical and iterative statistical methods, in addition to forward selection and backward elimination processes, were applied to identify explanatory variables. Regional random effect was checked, and there was no difference among regions.

## Results

### Characteristics of Health Facilities

A total of 131 health facilities (113 health centre and 18 hospitals) across 56 woredas, six zones, and three regions participated in the zonal-based assessment of organizational improvement factors (Fig. 2). The majority of facilities were health centers (113, 86.3%), followed by primary hospitals (11, 8.4%).

**Fig 2:**
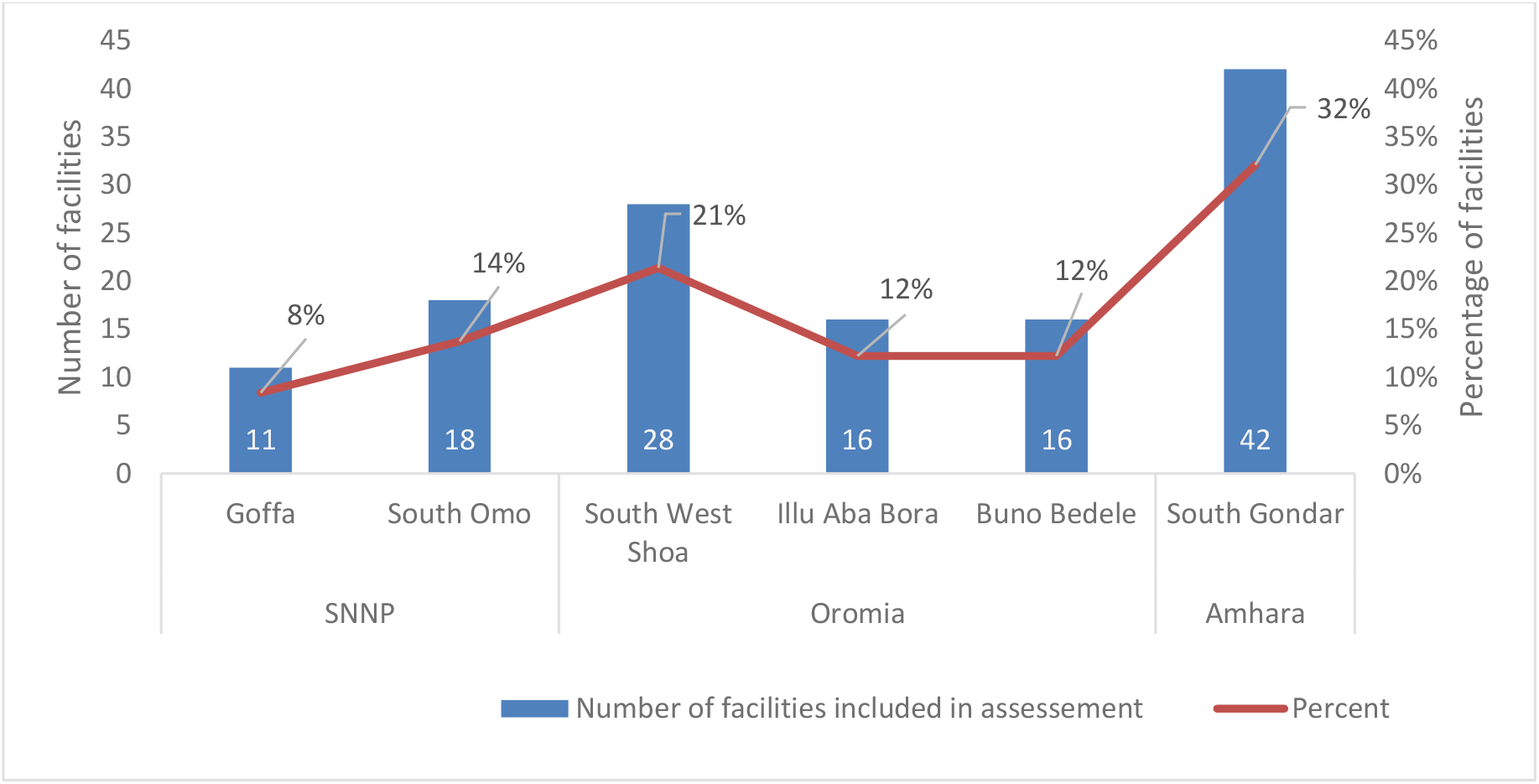
Number of health facilities included in the evaluation assessment (N=131)

Among these facilities 105 (80.2%) had grid electricity, and about half of them had a backup electricity supply. However, more than one-fourth of the health facilities lacked a water pipeline (Table 2).

**Table 2:** Characteristics of Health facilities participated in the study, May 2022, N=131.

| Variable | Responses | Health facilities (n, %) |
| --- | --- | --- |
| Facility Type | Health Centre | 113 (86.3) |
|  | Primary Hospital | 11 (8.3) |
|  | General Hospital | 5 (3.8) |
|  | Teaching Hospital | 2 (1.5) |
|  | Total | 131 (100.0) |
| Source of Power Supply | None | 2 (1.5) |
|  | Solar | 19 (14.5) |
|  | Generator | 5 (3.8) |
|  | Main Grid | 50 (38.2) |
|  | Main Grid with backup | 55 (42.0) |
|  | Total | 131 (100.0) |
| Source of Water Source | Pipeline | 96 (73.3) |
|  | Under Groundwater | 15 (11.5) |
|  | Roof collected water | 6 (4.6) |
|  | River water | 9 (6.9) |
|  | Other sources | 5 (3.8) |
|  | Total | 131 (100.0) |

### Organizational factors

#### Antecedents and Supports for Quality improvement

Fifty-nine percent of health facilities had two or fewer QI-trained personnel, and only 16% had five or more QI-trained staff. In most health facilities (75.6%), QI tools and methods were used more frequently beyond the first QI project design to address quality problems in routine practice. Fishbone, run chart, and prioritization matrix were the most commonly utilized QI tools by 90.8%, 76.3%, and 71.8% of the health facilities, respectively, as shown in Figure 3. IHI provided QI coaching to 77 (78.8%) health facilities, while coaching zonal and woreda QI coaches to build a sustainable QI cadre within the local system. Eighty-three-point two percent (83.2%) of health facilities received coaching from zonal and woreda QI coaches at least once per year, supporting the implementation of quality plans, dashboards, and improvement projects (Table 3).

**Fig 3::**
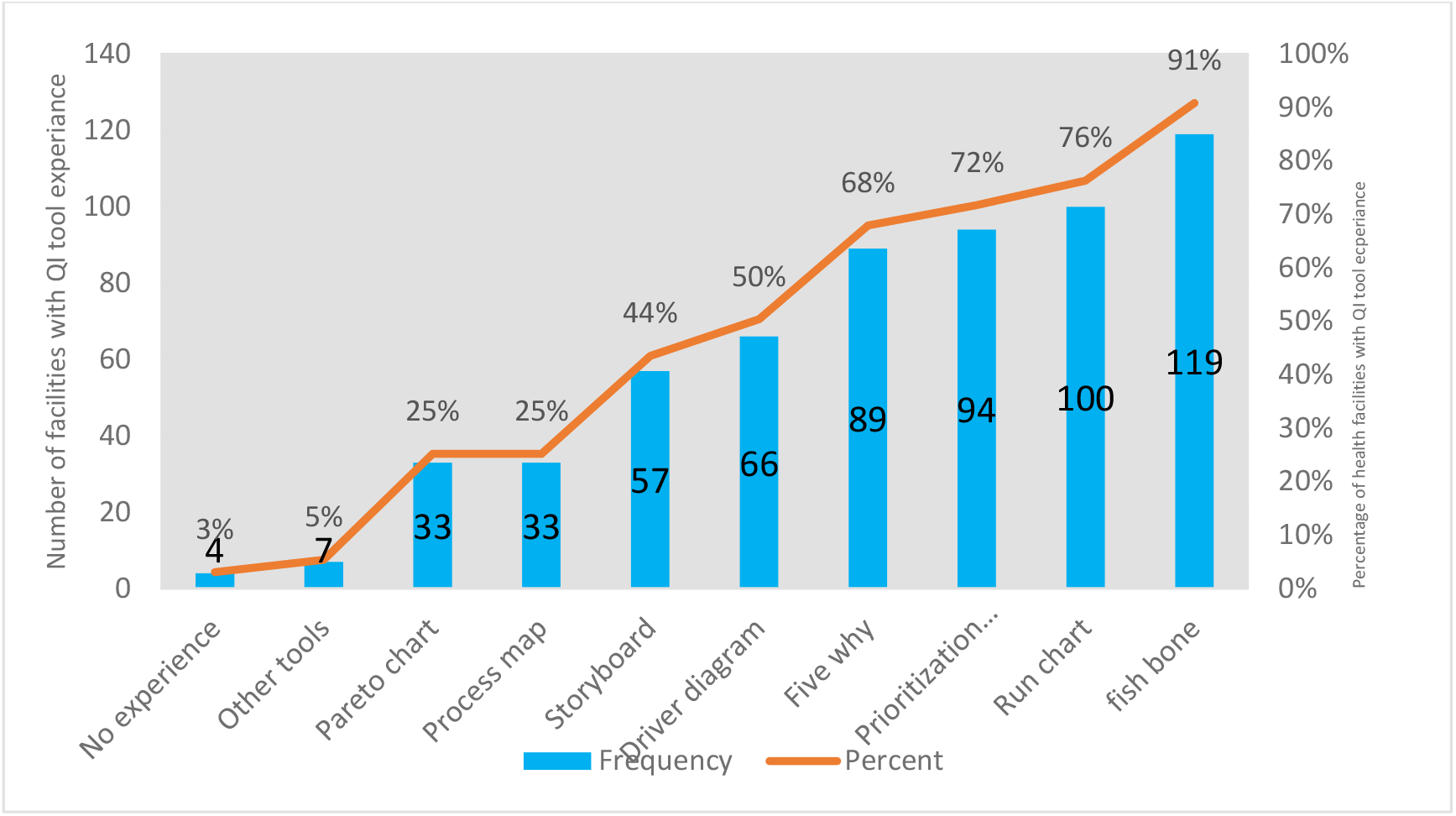
Health facilities’ experience of using QI tools and methods from April 2020-March 2022, N=131

#### Leadership engagement in Quality improvement

A quality plan was developed by 45% of health facilities for the year 2020/2021, and 42% had an updated quality plan for the year 2021/2022. Eighty-one (61.8%) health facilities had regular (i.e. at least once per month in the first six of the intervention) documented QI team meetings, and facility leaders participated in more than 90% of QI team meetings. One hundred two (77.9%) of health facility leaders used the QI dashboard to inform decision-making during QI or performance review meetings. The facility leaders participated in 86.7% health facilities’ QI project design process. In the majority of health facilities (70%), leaders assigned the required human resources to QITs to implement specific QI activities. The necessary equipment and supplies, and the budget for QI project implementation, were allocated to 59.5% and 30.5% of Health facilities, respectively (Table 4 and Fig 4).

**Fig 4:**
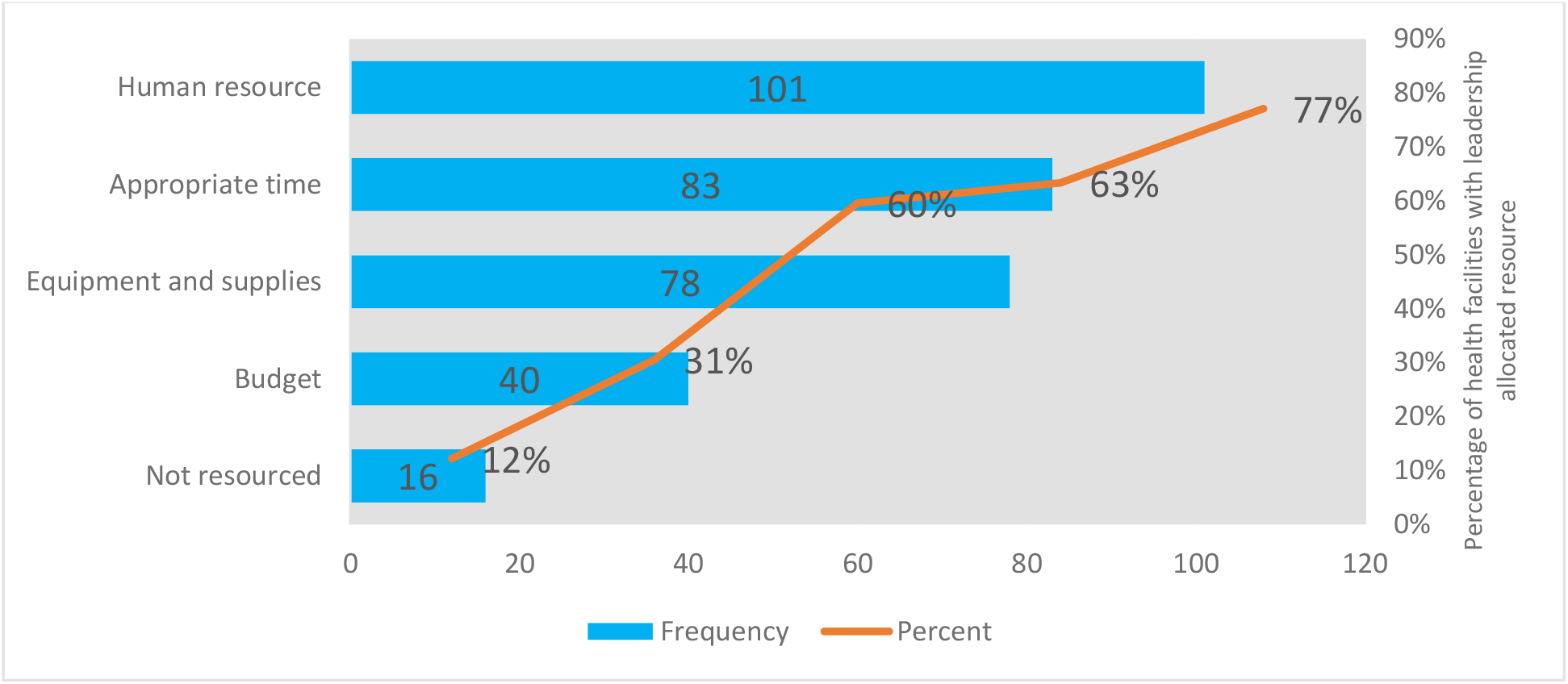
Health facilities with leadership allocated resources to achieve QI project aim April 2020-March 2022, N=131

#### Staff engagement in quality improvement

While all assessed health facilities had established quality structures with designated personnel, such as a focal person, quality unit, or quality directorate, the level of trained human resources within these structures varied. However, only 75 (57.3%) of health facilities had quality structures with transparent Terms of Reference (ToR). This means the ToR was jointly developed and shared with staff involved in QI activities. Staff directly involved in QI activities in 85.5% of health facilities received constructive feedback from their supervisors within the year. However, staff recognition mechanisms for QI achievements were limited. Only 40 (30.5%) health facilities acknowledged facility staff for their accomplishments in QI implementation (Table 5).

#### Health facilities readiness for Quality improvement

Health facilities readiness for QI scale-up implementation includes conducting baseline assessments by the facilities themselves, having quality data, incorporating QI activities into staff job descriptions, community feedback receiving platforms, and participating in learning sessions. All health facilities assessed in this evaluation conducted baseline assessment at least for the targeted MNH bundle and MNH quality indicators, and identified major QI priority areas. The majority (87%) of health facilities maintained reporting timeliness, and more than 84% documented an average annual LQAS score above 90% in the year before data collection. Thirty-one percent of health facilities incorporated QI activities into their staff’s job descriptions as an organizational strategy to strengthen ownership and sustainability of QI implementation beyond trained personnel. Health facilities had the opportunity to attend QI-related learning sessions or review meetings at different levels, accounting for 64.1% of the total, as shown in Table 6. All health facilities had at least one mechanism for receiving community feedback. The assessment found that many health facilities (71.8%) used feedback boxes to receive community reflections (Table 6 and Fig 5).

**Fig 5:**
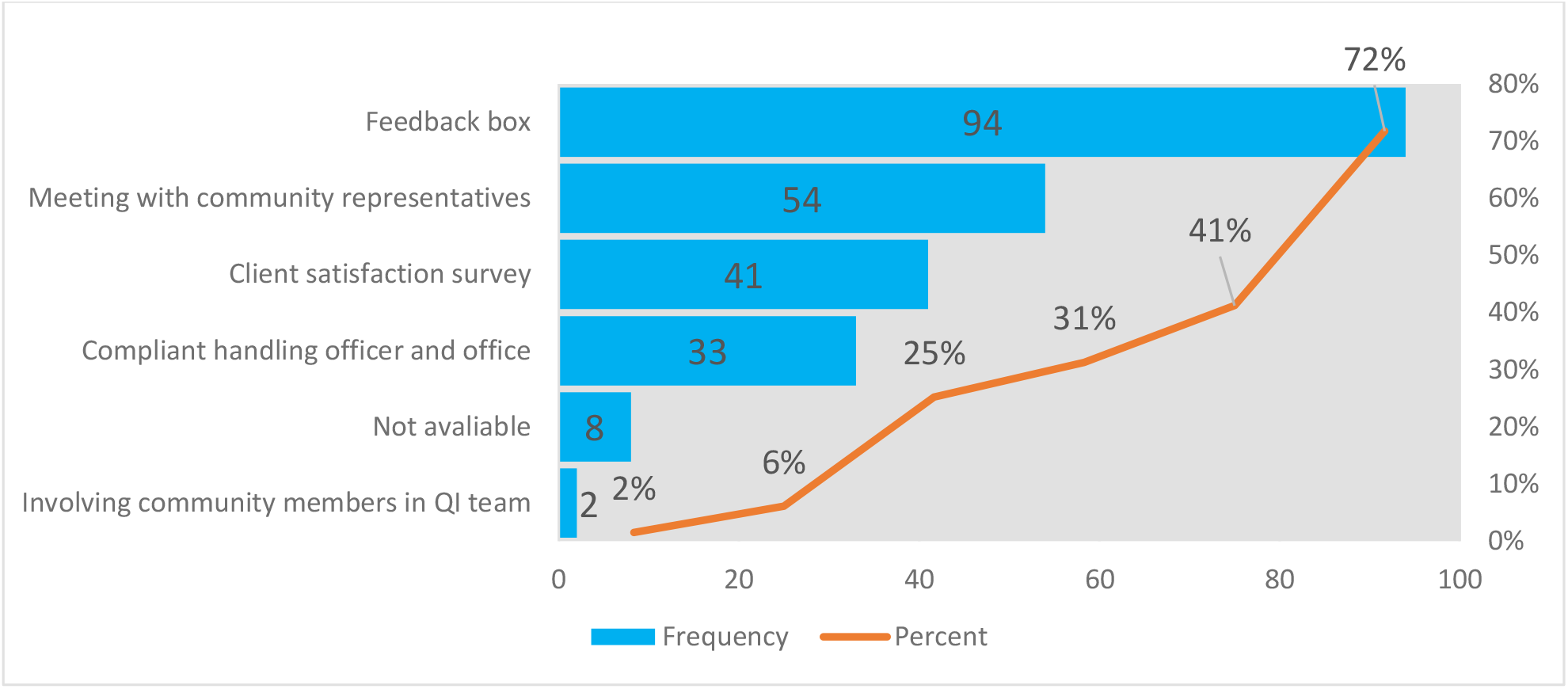
Health facilities client feedback mechanisms for Quality improvement, April 2020-March 2022, N=131

#### Quality improvement supportive activities

Health facilities begin QI scale-up by providing on-site orientation to QI methods and tools for their staff, using formally trained QI team members. In this evaluation, most health facilities (107, 81.7%) provided staff orientation to an average of 13.6 staff members before designing and implementing QI projects. Each health facility engaged an average of 10 staff members in designing and implementing QI projects. More than half (67/51.1%) of health facilities conducted clinical audits every quarter or every six months as part of QI implementation within their respective facilities (Table 7).

#### Quality initiative scale-up implementation achievements

Quality improvement practice were assessed using the clinical bundle measures and the MNH quality indicators. Overall, 103 (78.6%) health facilities had at least one improvement in a clinical bundle indicator, based on run chart rules. Similarly, 117 (89.3%) facilities showed improvements in at least one MNH quality indicator (Table 8).

**Table 8:** Improved Clinical Bundle and HIMS Indicators by each health facility from April 2020 to March 2022, N=131.

| Number of improved Clinical bundle indicators | Health facilities (n, %) | Comm. | Number of improved MNH quality indicators | Health facilities (n, %) | Comm. |
| --- | --- | --- | --- | --- | --- |
| 0 | 28 (21.37) | 21.37 | 0 | 14 (10.69) | 10.69 |
| 1 | 32 (24.43) | 45.80 | 1 | 29 (22.14) | 32.82 |
| 2 | 25 (19.08) | 64.89 | 2 | 31 (23.66) | 56.49 |
| 3 | 21 (16.03) | 80.92 | 3 | 34 (25.95) | 82.44 |
| 4+ | 25 (19.09) | 98.47 | 4 | 13 (9.92) | 92.37 |
| Total | 131 (100.0) |  | 5+ | 10 (7.64) | 100 |

### Factors associated with Improved Clinical Bundle and MNH quality indicators

#### Factors associated with improved clinical bundle indicators

As shown in Table 9, the mixed-effect Possin regression analysed the impact of organizational factors on improved clinical bundle indicators across the three regions. The overall model was statistically significant, with an incidence rate ratio for improved indicators of 1.82 (95% CI [1.11, 2.99]). The exploratory analysis revealed that health facilities in which respondents (facility managers or designated key informants) perceived teamwork as a critical factor had a higher number of improved bundle indicators compared to those where teamwork was not perceived as critical factor (AIRR = 1.87; 95% CI [1.06, 3.29]). Similarly, health facilities that conducted client satisfaction surveys had an incident rate for improved clinical bundle indicators of 1.27 (95% CI [1.12, 1.44]) times that of health facilities that couldn’t conduct client satisfaction surveys. Moreover, health facilities with a quality structure and clear terms of reference (ToR) had an incident rate of 1.34 (95% CI [1.23, 1.47]) times that of facilities without a quality structure with clear ToR.

**Table 9:**
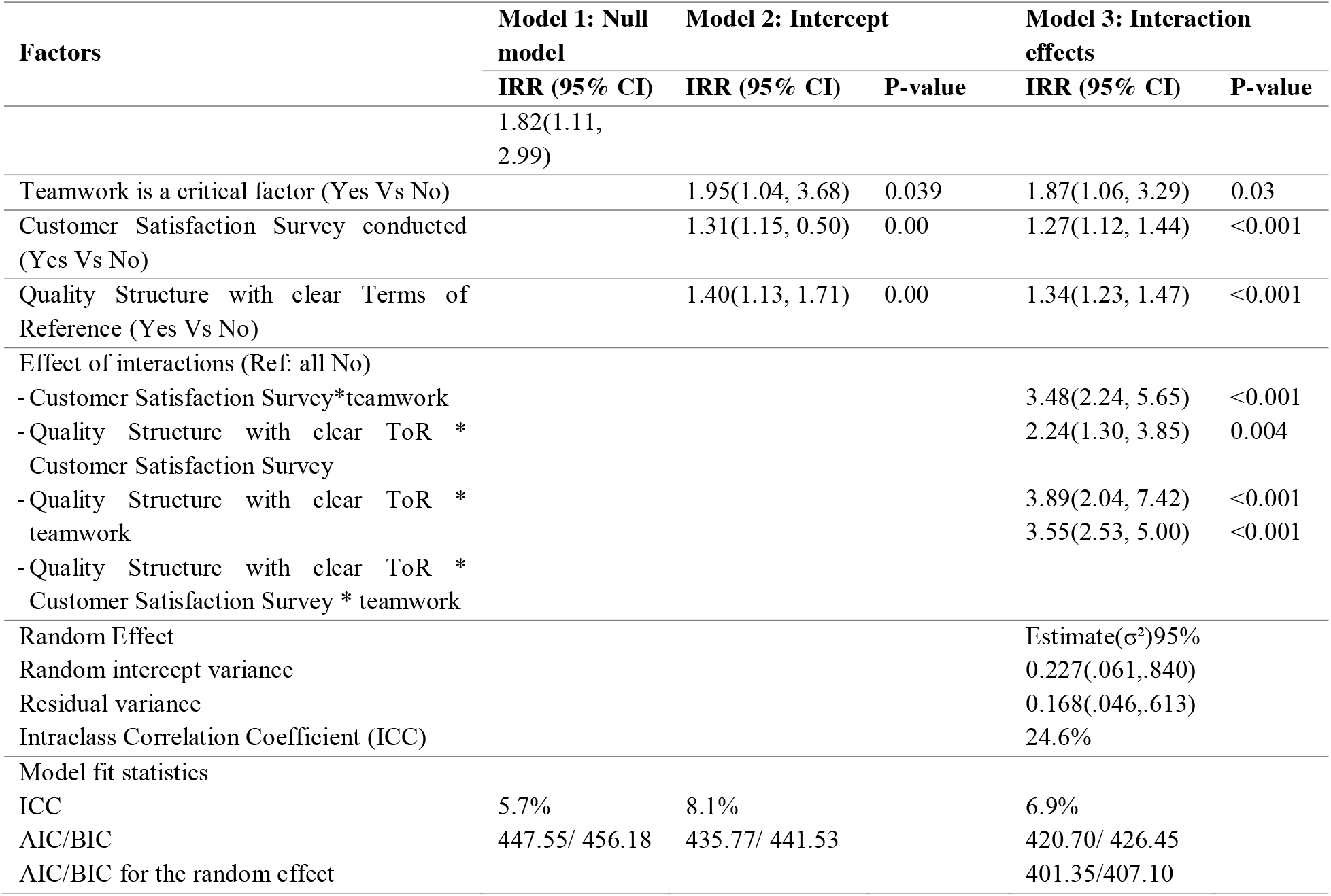
Mixed-effect Poisson regression showing factors associated with the number of improved clinical bundles, April 2020 to March 2022, N=131.

| Factors | Model 1: Null model | Model 2: Intercept |  | Model 3: Interaction effects |  |
| --- | --- | --- | --- | --- | --- |
|  | IRR (95% CI) | IRR (95% CI) | P-value | IRR (95% CI) | P-value |
|  | 1.82(1.11, 2.99) |  |  |  |  |
| Teamwork is a critical factor (Yes Vs No) |  | 1.95(1.04, 3.68) | 0.039 | 1.87(1.06, 3.29) | 0.03 |
| Customer Satisfaction Survey conducted (Yes Vs No) |  | 1.31(1.15, 0.50) | 0.00 | 1.27(1.12, 1.44) | <0.001 |
| Quality Structure with clear Terms of Reference (Yes Vs No) |  | 1.40(1.13, 1.71) | 0.00 | 1.34(1.23, 1.47) | <0.001 |
| Effect of interactions (Ref: all No) |  |  |  |  |  |
| - Customer Satisfaction Survey*teamwork |  |  |  | 3.48(2.24, 5.65) | <0.001 |
| - Quality Structure with clear ToR * Customer Satisfaction Survey |  |  |  | 2.24(1.30, 3.85) | 0.004 |
| - Quality Structure with clear ToR * teamwork |  |  |  | 3.89(2.04, 7.42) | <0.001 |
| - Quality Structure with clear ToR * Customer Satisfaction Survey * teamwork |  |  |  | 3.55(2.53, 5.00) | <0.001 |
| Random Effect | | | | Estimate($\sigma^2$)95% | |
| Random intercept variance |  |  |  | 0.227(.061,.840) |  |
| Residual variance |  |  |  | 0.168(.046,.613) |  |
| Intraclass Correlation Coefficient (ICC) |  |  |  | 24.6% |  |
| Model fit statistics |  |  |  |  |  |
| ICC | 5.7% | 8.1% |  | 6.9% |  |
| AIC/BIC | 447.55/ 456.18 | 435.77/ 441.53 |  | 420.70/ 426.45 |  |
| AIC/BIC for the random effect |  |  |  | 401.35/407.10 |  |

The cross-level interaction model, Model 3, health facilities implementing all three interventions (customer survey*teamwork*quality structure with clear ToR) showed the strongest effects of 3.55 (95% CI [2.53, 5.00]) times better outcomes versus facilities with none of them. Those health facilities with effect of interactions between customer satisfaction survey*teamwork, quality structure with clear ToR*customer satisfaction survey and quality structure with clear ToR*teamwork had 3.48 (95% CI [2.24, 5.65]), 2.24 (95% CI [1.30, 3.85]) and 3.89 (95% CI [2.04, 7.42]) times better outcomes than those health facilities with neither of them, respectively (Table 9 and Fig 6).

**Figure 6:**
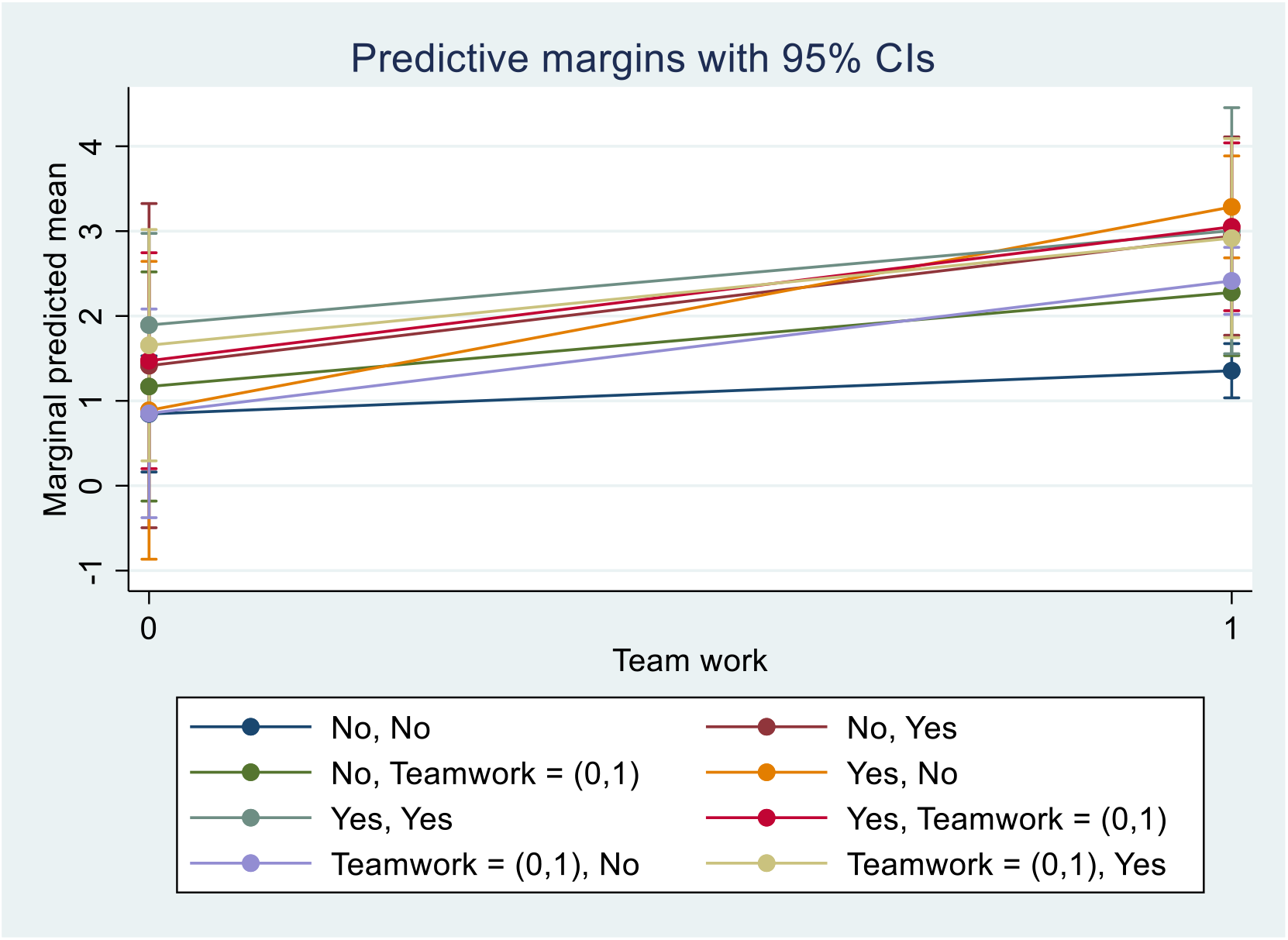
Marginal effects of improved clinical bundle indicators on teamwork across quality structure and customer satisfaction survey

The random effect showed that regional variability in both the baseline outcome (random intercept) and the effect of interventions, such as teamwork, was significant. The total intraclass correlation coefficient (ICC) was 24.6% with the random intercept variance of 0.227 (95% CI [0.061, 0.840]) and residual (facility level intervention) variance of 0.168 (95% CI [0.046, 0.613]) (Table 9).

#### Factors associated with improved MNH quality indicators

As shown in Table 10, a Possin regression was used to analyse the impact of organizational factors on improved MNH quality indicators across the three regions. The overall model was statistically significant, indicating that the predictors significantly impacted the outcome. The incidence rate for improved indicators was 2.31 times higher (95% CI [2.06, 2.58]), suggesting a strong association between the predictors and the likelihood of improving indicators. The exploratory analysis revealed that health facilities with knowledge of quality improvement (QI) had 1.34 times better outcomes (95% CI [1.08, 1.67]) than those that didn’t. Similarly, hospitals had 1.35 times better (95% CI [1.08, 1.67]) improved MNH quality indicators than health centers. On the other hand, those health facilities that reported resistance to change (AIRR = 0.73 (95% [0.58, 0.92]) and staff turnover (AIRR = 0.62 (95% [0.50, 0.76]) were less likely to achieve a better outcome.

**Table 10:** Poisson regression analysis showing factors associated with the number of improved MNH quality indicators, April 2020 to March 2022, N=131.

| Factors | Model 1: Null model | Model 2: Univariate regression model (crude) |  | Model 3: multivariable model (adjusted) |  |
| --- | --- | --- | --- | --- | --- |
|  | IRR (95% CI) | IRR (95% CI) | P-value | IRR (95% CI) | P-value |
|  | 2.31 (2.06, 2.58) |  |  |  |  |
| Health Facility Type (Ref: health center) |  |  |  |  |  |
| Hospital |  | 1.79 (1.39, 2.31) | <0.001 | 1.35(1.04, 1.74) | 0.024 |
| Knowledge-QI (Ref: Not a critical factor) |  |  |  |  |  |
| Yes, it is a critical factor |  | 1.51(1.19, 1.91) | 0.001 | 1.34(1.08, 1.67) | 0.009 |
| Resistance to change (Yes vs No) |  | 0.67(0.51, 0.86) | 0.002 | 0.73(0.58, 0.92) | 0.007 |
| Staff Turn Over (Yes vs No) |  | 0.58(0.46, 0.73) | <0.001 | 0.62(0.50, 0.76) | <0.001 |
| Staff satisfaction assessment in the health facility (Yes vs No) |  | 1.61(1.27, 2.05) | <0.001 | 1.20(0.83, 1.74) | 0.343 |
| Staff recognition mechanisms for their QI activities achievement (Yes vs No) |  | 1.30(1.03, 1.65) | 0.027 | 1.15(0.95, 1.38) | 0.164 |
| Structure Quality existence in the facility (Yes vs No) |  | 1.30(1.01, 1.66) | 0.043 | 0.74(0.50, 1.10) | 0.135 |
| Leadership Support (Ref: Not a critical factor) |  |  |  |  |  |
| Yes, it is a critical factor |  | 1.41(1.03, 1.95) | 0.034 | 1.04(0.78, 1.39) | 0.795 |
| **Model fit statistics** |  |  |  |  |  |
| AIC/BIC | 470.99/473.87 | 461.87/467.62 |  | 438.52/452.90 |  |

## Discussion

Based on the set indicators assessment, the post-assessment showed that most health facilities (about 89%) and MNH indicators (about 79%) had at least one indicator improvement in the clinical bundle and MNH indicators, respectively. This suggests that the quality initiative scale-up project was associated with measurable improvements in selected indicators across facilities. However, these improvements should be interpreted in relation to baseline gaps, facility priorities, and implementation context. A project evaluation study supports similar findings, although differences in intervention design and geographical settings may explain variation in performance levels (24-27).

The study revealed variation between regions. A meaningful proportion of variability in outcomes was attributable to regional clustering, as reflected in the ICC of 24.6%, while most variability was attributable to differences between individual health facilities (75.4%). This implies the presence of contextual (cluster-level) effects, although facility-level characteristics appeared to play a predominant role (28-31).

In this study, associated factors were identified with the incidence of improved clinical bundle and MNH quality indicators. For improved clinical bundle indicators, significant association were observed with perceived teamwork, customer satisfaction survey and the presence of a quality structure with clear ToR. For improved MNH quality indicators, significant association were observed with health facility type, knowledge about QI, resistance to change and staff turnover (5, 32). Teamwork was perceived as a critical factor associated with improved clinical bundle performance. This reflects the role of multidisciplinary collaboration in integrating knowledge, skill, experience, and perspectives support effective problem-solving and sustained improvement (23, 33).

The presence of a quality structure with a clear ToR was significantly associated with improved clinical bundle measures. This suggests that formal organizational arrangements may support accountability, coordination, and effective implementation of QI activities. Evidence from Alexander *et al*. supports the importance of organizational infrastructure in facilitating quality improvement and improving health outcomes (34).

The study revealed evidence of effect modification among teamwork, client satisfaction survey, and the presence of a quality structure with a clear ToR in relation to improved clinical bundle indicators. Interaction effects suggest a synergistic relationship between these factors, with stronger effects observed when combined. Health facilities implementing all three components (customer survey ^x^ teamwork ^x^ quality structure with clear ToR) showed the strongest association, with outcomes 3.55 times higher (95% CI [2.53, 5.00]) compared to facilities without these components. This suggests that combined implementation of multiple QI components may produce greater improvements than single intervention alone (35).

On the other hand, knowledge of quality improvement was significant factor associated with improved MNH quality indicators. Batalden and Stoltz emphasize that combining professional with improvement knowledge is essential for sustained enhancement of healthcare outcomes (36). Staff resistance was another significant factor associated with improved MNH quality outcomes. This may reflect challenges such as a limited understanding the improvement objectives and reluctance to change established practices. Although adaptation takes time, engaging sceptical staff can facilitate their transition into active contributors to improvement efforts. A qualitative study by R Gollop et al., highlights that health worker engagement, particularly among physicians, is essential for sustaining quality improvement initiatives (37). Similarly, Wilcock and Lewis provide evidence that continuous quality improvement approaches can support sustained change and skill development over time (38).

Staff turnover was also significantly associated with improved MNH quality indicators. This finding highlights the importance of workforce stability and sustained skill retention in supporting quality improvement efforts. Studies have shown that higher staff continuity is associated with improved treatment quality and organizational performance (36). Staff turnover has also been linked to Staff turnover has also been linked to reduced morale, productivity, and innovation, underscoring the need for effective retention strategies to support patient outcomes (39, 40).

It is also worth noting that several QI process factors, including but not limited to staff training, leadership engagement, use of QI tools, coaching support, and data use practices, were included in the multivariable analysis but were not statistically significant predictors of outcome indicators. Given that the unit of analysis was health facilities (n = 131), this may reflect limited statistical power as well as variability in the intensity and consistency of implementation across facilities. Although not statistically significant in adjusted models, the descriptive findings suggest that outcome improvements occurred within a broader context of QI system strengthening. These activities formed part of the implementation package delivered during scale-up and represent core processes through which facilities engaged in quality improvement practice.

## Strengths and Limitations

This study has several strengths. It included health facilities across three major regions of the country. Multiple data sources were used, including interviews, charts, register reviews, and electronic reports. Data were collected by senior experts in healthcare quality, data collection, and processing. Additionality, the use of a mixed Poisson model accounted for variability across regions, further strengthening the validity of the findings.

The study also had limitations. Direct observation was not applied, and reliance on medical chart reviews may have introduced data validity issues in assessing improved clinical bundle indicators. Furthermore, population projections were used to estimate the denominators for MNH quality indicators, which may have affected the accuracy of the results. In addition, not all potential facility or individual-level factors influencing QI implementation (such as frontline staff perspectives, motivation, and experience) were captured in the quantitative analysis, resulting in possible residual confounding.

## Implications for Practice and Policy

Our findings have several important implications for improving maternal and newborn healthcare quality in low-resource settings:

Strengthening teamwork and leadership engagement: Collaborative working environments, supported by active facility leadership, are critical for the consistent implementation of care standards. Investing in team cohesion and leadership engagement may support more effective of quality improvement implementation.

Integrated organizational interventions: Quality improvement interventions appear more effective when implemented as integrated approaches, combining teamwork, formal quality governance structures, and routine client feedback, rather than as isolated activities.

Institutionalizing quality improvement systems: Sustaining improvements requires the institutionalization of QI system alongside ongoing capacity building for staff. Clearly defined roles within quality teams, structured support for problem-solving, and continuous learning mechanisms may contribute to sustained implementation over time.

Workforce stability and change management: High staff turnover and resistance to change can undermine quality improvement efforts. Strategies to enhance workforce stability, staff engagement, and readiness for change including retention approaches and structured change management may help sustain and scale improvements.

Overall, these findings highlight the importance of organizational and system-level approaches alongside clinical interventions in improving maternal and newborn healthcare outcomes. Addressing both structural and human factors may support stronger health systems and more sustainable quality improvement efforts.

## Conclusion

The study found that most health facilities demonstrated improvement in at least one clinical bundle and MNH quality indicator. Several factors were significantly associated with improvements in clinical bundle indicators, including teamwork, the use of client satisfaction surveys, and the presence of a facility quality structure with clear terms of reference. For MNH quality indicators, significant associations were observed with health facility type, knowledge of quality improvement, reported staff resistance to change, and staff turnover.

These findings suggest that organizational factors may play an important role in supporting quality improvement efforts. Accordingly, program implementers may consider giving attention to these factors when designing and implementing quality initiatives. Given that the study relied on historical records, further research using observational or longitudinal designs would help to strengthen the evidence base.

## Data Availability

Data is available upon request.

## List of abbreviations

ANC: Antenatal care
IHI: Institute for Healthcare Improvement
MNH: Health Management Information System
MNH: Maternal and neonatal health
QI: Quality improvement
SNNP: South Nations, Nationalities and Peoples
ToR: Terms of References
WoHO: Woreda health offices
ZHD: Zonal health departments

## Declarations

### Ethics approval and consent to participate

Ethical approval for the evaluation was obtained from the Ethiopian Public Health Association (EPHA) Scientific and Ethical Review Committee (Ref: EPHA/OG/129/21). As this study was conducted as part of a program evaluation using chart reviews and routine project data, the requirement for informed consent was waived by the IRB in accordance with the Ethiopian ethical guidelines (41). A support letter was obtained from relevant health administrative bodies, and permission was secured from the respective health facilities before data collection. The purpose and importance of the evaluation were explained to all participants, and participation was entirely voluntary. Written informed consent was obtained from all participants before primary data collection, with each participant providing a signed consent form approved by the IRB. For medical record reviews, informed permission was obtained from health facility heads on behalf of the mothers, as direct contact with the clients was not feasible. Confidentiality of the information was strictly maintained by omitting personal identifiers, and the data were used solely for research purposes.

### Consent for publication

Not applicable.

### Availability of data and materials

https://zenodo.org/records/17055840

### Author Contributions

NW, ABK and AK developed the study protocol. NW and AK conceptualized the study, contributed to the development of the research questions and reviewed data analytical methods. NW and GT analysed the descriptive data and contributed to manuscript writing. ABK did the full data analysis and wrote the draft manuscript. NW, GT, AA, MA and AK reviewed and provided substantial revisions to the draft manuscript. All authors reviewed and approved the final version of the manuscript.

## Acknowledgements

We would like to extend our sincere thanks to the Ethiopian Ministry of Health, regional health bureaus, zonal health departments, woreda health offices, and health facility staff for their unreserved support and collaboration throughout the data collection process. We thank the data collectors for gathering the data that informed this analysis.

## Supplementary file

Additional file 1: A conceptual framework, entitles as “S1 Conceptual Framework”.

Additional file 2: Sample Run charts, which are used for improved indicators, are entitled “S2 Sample run charts”.

Additional file 3: List of indicators used for outcome measurement for this paper, entitled as “S3 List of indicators and bundle elements”.

Additional file 4: English version of the questionnaire entitled: “S4 Facility Questioner Organization level 4th”.

